# Hyperactive immature state and differential CXCR2 expression of neutrophils in severe COVID-19

**DOI:** 10.1101/2022.03.23.22272828

**Authors:** Christopher M. Rice, Philip Lewis, Fernando M. Ponce-Garcia, Willem Gibbs, Drinalda Cela, Fergus Hamilton, David Arnold, Catherine Hyams, Elizabeth Oliver, Rachael Barr, Anu Goenka, Andrew Davidson, Linda Wooldridge, Adam Finn, Laura Rivino, Borko Amulic

## Abstract

Neutrophils are vital in defence against pathogens but excessive neutrophil activity can lead to tissue damage and promote acute respiratory distress syndrome (ARDS). COVID-19 is associated with systemic expansion of immature neutrophils but the functional consequences of this shift to immaturity are not understood. We used flow cytometry to investigate activity and phenotypic diversity of circulating neutrophils in acute and convalescent COVID-19 patients. First, we demonstrate hyperactivation of immature CD10^−^ subpopulations in severe disease, with elevated markers of secondary granule release. Partially activated immature neutrophils were detectable three months post symptom onset, indication long term myeloid dysregulation in convalescent COVID-19 patients. Second, we demonstrate that neutrophils from moderately ill patients downregulate the chemokine receptor CXCR2, while neutrophils from severely ill individuals failed to do so, suggesting altered ability for organ trafficking and a potential mechanism for induction of disease tolerance. CD10^−^and CXCR2^hi^ neutrophil subpopulations were enriched in severe disease and may represent prognostic biomarkers for identification of individuals at high risk of progressing to severe COVID-19.

## Introduction

Severe acute respiratory syndrome coronavirus 2 (SARS-Cov2), the causative agent of COVID-19, is a single stranded RNA virus that is transmitted by respiratory droplets. The clinical spectrum of COVID-19 is wide, ranging from a paucisymptomatic self-limiting upper airway infection to a lower respiratory tract infection (1), associated with acute respiratory distress syndrome (ARDS). The hyper inflammatory state in ARDS impairs lung function and is the cause of death in 70% of such patients (1, 2). Other systemic manifestations of COVID-19 include thromboembolism (3), acute kidney damage (4) and cardiac injury (5).

Excessive host inflammatory responses are recognized as central to the pathogenesis of severe COVID-19. Viral proliferation induces type I interferons and proinflammatory cytokines (6), which may be protective early in infection, but lead to cytokine storm and tissue damage when produced excessively (7). Immunosuppressive therapies, such as the glucocorticoid dexamethasone, significantly reduce mortality in hospitalized patients receiving respiratory support (8). Dexamethasone therapy, however, is not successful in all patients and the mechanistic basis for this failure is not understood.

Neutrophils are abundant myeloid cells that are vital for defense against pathogens, acting to suppress bacterial and fungal dissemination. Their antimicrobial response includes production of reactive oxygen species (ROS), degranulation of proteases and release of extracellular chromatin in the form of neutrophil extracellular traps (NETs) (9). The role of neutrophils in the anti-viral response is less clear (10). Importantly, excessive or dysregulated neutrophil activity can promote immunopathology and is thought to be a key feature of ARDS (11).

Despite improved insight into the inflammatory injury in SARS-CoV-2 infection, our understanding of the neutrophil contribution remains incomplete. Transcriptomic and proteomic analyses of COVID-19 patient peripheral blood (12-16) and bronchoalveolar lavage fluid (17, 18) reveal a disordered myeloid response. Peripheral neutrophil activation predicts clinical outcome and is strongly associated with mortality (12, 13, 15). Similarly, circulating neutrophil counts are elevated in severely ill patients and higher neutrophil-to-lymphocyte ratios are associated with poor prognosis (19, 20). Neutrophil chemoattractants, such as CXCL1 and CXCL8 (IL-8) (21, 22), as well as transmigrated, infiltrating neutrophils (21, 23-25) are detected in the alveolar space of infected lungs. Neutrophil soluble mediators, such as elastase and calprotectin, accumulate in plasma of patients with severe disease (26, 27). Furthermore, NETs are elevated in plasma of severe patients and correlate with lung failure and immunothrombosis (23, 28-30). Several studies have also reported expansion of low density (31) and immature neutrophils (27, 32-34), whose role in disease remains unclear. Collectively, these observations suggest that neutrophil activation plays a significant role in disease progression and eventual fatality of COVID-19.

We performed a detailed multidimensional flow cytometry analysis and proteomic characterisation of circulating neutrophils in patients with COVID-19. We discerned neutrophils with high discrimination from closely related leukocyte subsets and examined receptors associated with activation, maturation and trafficking. COVID-19 patients exhibit a pattern of systemic neutrophil activation and increased neutrophil heterogeneity. Hyperactive immature states and maintenance of CXCR2 are prognostic of poor patient outcome. Our study highlights the importance of neutrophil developmental state when considering pathogenic function in COVID-19 and identifies potential biomarkers of severe disease.

## Results

### Immature circulating neutrophils are enriched in moderate and severe COVID-19

To assess phenotypic changes in neutrophil populations, we analysed fresh peripheral blood from hospitalised acute COVID-19 patients (n=34, median days since symptom onset = 13±5.29; median days since hospital admission = 4±3.99), convalescent individuals (n=38, days since symptom onset median=76.5±30.7) and healthy controls (n=20) (Table 1). Patients were stratified by disease severity (35), with severe disease defined by requirement for ventilation and/or intensive care admission, moderate disease defined as requirement for supplementary oxygen without intensive care and mild disease requiring no oxygen supplementation and no intensive care. Stratification of patients was representative of status at time of sample acquisition, except when analysing survival. Patients with severe disease were further stratified into surviving and deceased groups.

**Table 1:** Cumulative clinical data of COVID-19 patients. * Data not available ** patients did not report a symptom onset date.

COVID-19 patients
|  | Mild (n=3) | Moderate (n=14) | Severe (n=18) |
| --- | --- | --- | --- |
| **Demographic and clinical data** |  |  |  |
| Age, years median ± standard deviation | 78±6.48 | 52.5±10.59 | 65±13.31 |
| Sex, M/F | 0/3 | 9/5 | 14/4 |
| Intensive/high care admission (%) | 0% | 7% | 83% |
| Deceased within 33 days of admission | 0/3 | 0/14 | 6/18 |
| **Peripheral Blood Laboratory Parameters**<br>median ± standard deviation | | | |
| Leukocyte count $\times 10^9/L$ | 6.36±1.4 | 6.74±3.7 | 8.69±5 |
| Neutrophil count $\times 10^9/L$ | 4.59±0.89 | 4.79±3.1 | 7.35±4.79 |
| Lymphocyte count $\times 10^9/L$ | 1.22±0.42 | 1.09±0.47 | 0.95±0.49 |
| Neutrophil to lymphocyte ratio | 3.76±1.22 | 5.02±4.27 | 5.91±8.03 |
| Serum D-dimer | * | 0.59±6.03 | 0.56±0.12 |
| Serum CRP (mg/dL) | 42±91.68 | 62.5±45.3 | 131±109.4 |
| **Comorbidities** |  |  |  |
| Diabetes | 1/3 | 3/14 | 3/18 |
| Heart disease | 2/3 | 1/14 | 6/18 |
| Chronic kidney disease | 1/3 | 0/14 | 0/18 |
| **Sample timeline**<br>median ± standard deviation | | | |
| Days since symptom onset | ** | 9.5±4.45 | 14±5.07 |
| Days since hospitalisation | 5±5.1 | 2±1.87 | 5.5±4.07 |

Whole blood was labelled using a panel of antibodies targeting surface markers designed to identify neutrophils with a high degree of discrimination and assess phenotypic changes. We identified neutrophils as CD14^-^, CD15^+^, CD125^-^ (Sup Fig 1A), thereby avoiding the commonly used marker CD16 (FcϒRIII), which varies in expression with neutrophil maturity (36) and is the target of surface proteases (27, 37). To ensure CD15^+^ CD16^-^ eosinophils were not incorrectly identified as neutrophils, we excluded these based on of IL-5R (CD125) expression (Sup Fig 1A). Despite a trend towards an increased neutrophil count (Sup Fig 1B), and elevated neutrophil-to-lymphocyte ratio (Sup Fig 1C), in severely affected patients, no significant differences in these parameters were observed in our cohort.

CD10, also known as membrane metalloendopeptidase (MME), is a neutrophil maturity marker whose expression is absent or less abundant on immature neutrophils (38, 39). We detected reduced CD10 expression on neutrophils from moderate and severe COVID-19 patients compared to both healthy controls and convalescent individuals (Sup Fig 2A). Moderate and severe groups were also enriched in CD10 negative (CD10^−^) neutrophils (Fig 1A), determined by comparison to CD10^−^ monocytes (Sup Fig 2B). CD10 surface expression was lowest amongst patients that subsequently succumbed to the disease (non-survivors) (Sup Fig 2C), suggesting prognostic utility. We confirmed neutrophil immaturity by quantifying expression of an additional marker, CD101 (Sup Fig 2D), which is similarly reduced or absent on immature cells (36, 40). Taken together, our results conclusively demonstrate accumulation of immature neutrophils in circulation of COVID-19 patients, consistent with other reports (32, 41, 42).

**Figure 1:**
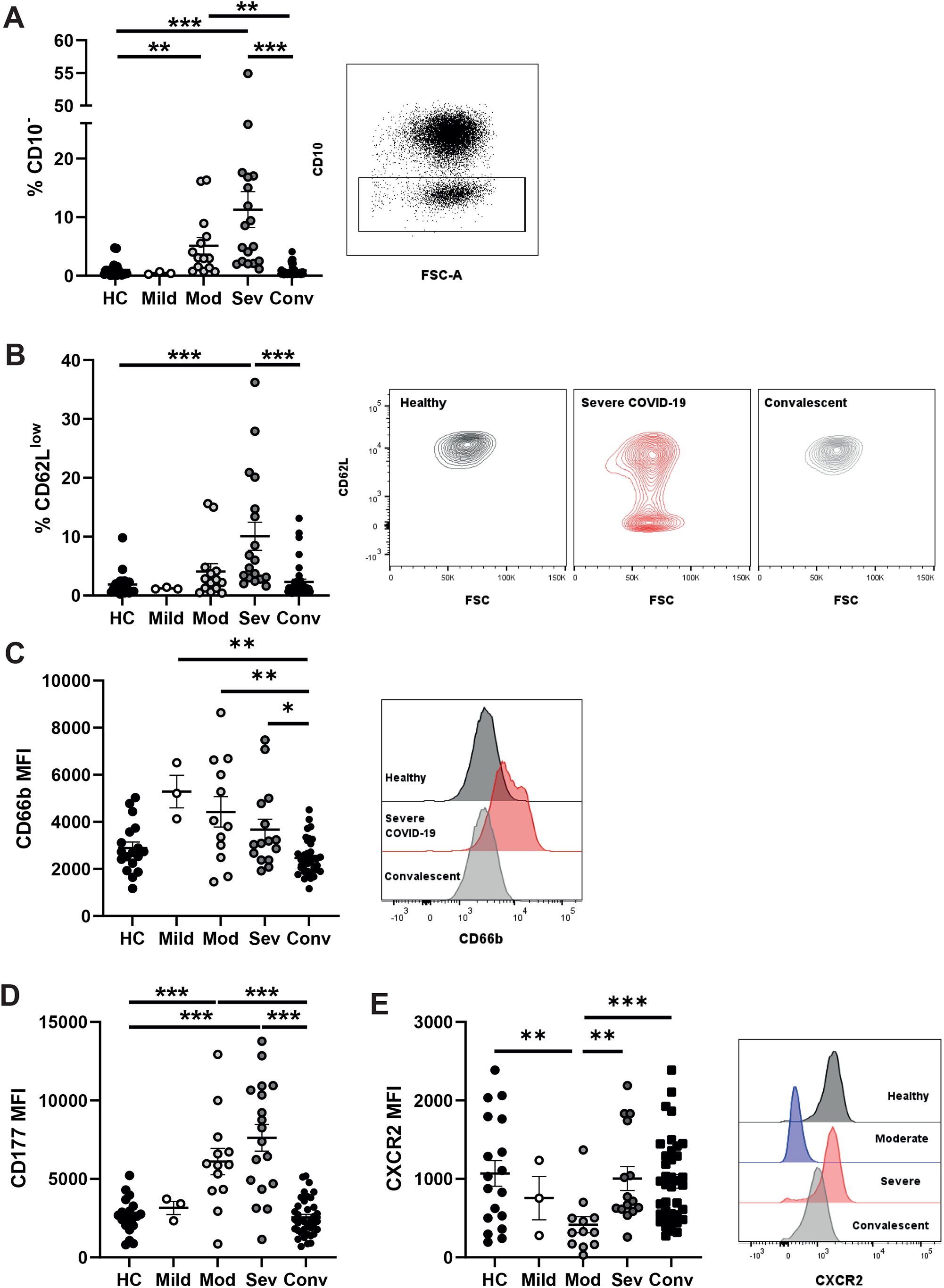
Alterations to circulating neutrophil phenotype during acute SARS-CoV-2 infection. Peripheral blood from healthy controls, convalescent and COVID-19 patients was analysed by flow cytometry. A) Percentage of CD10^−^ neutrophils from stratified groups and representative gating for CD10^−^ neutrophils. B) Percentage of CD62L^low^ neutrophils and representative contour plots of CD62L expression. C) Neutrophil CD66b median fluorescent intensity and representative histogram of CD66b fluorescent intensity. D) CD177 median fluorescent intensity on CD177^+^ neutrophils. E) Neutrophil CXCR2 median fluorescent intensity and representative histogram of CXCR2 fluorescent intensity. In A and B, data were analysed by Kruskal-Wallis test with Dunn’s multiple comparisons displayed on graph; in C, D and E, data were log transformed and analysed by one-way ANOVAs with Tukey’s multiple comparisons displayed on the graph. Values represent mean ± standard deviation.

### Peripheral neutrophil activation in severe COVID-19

We assessed neutrophil activity by quantifying reduction of CD62L (L-selectin) expression, a surface receptor that is proteolytically shed upon stimulation (43). CD62L^low^ neutrophils were elevated in severe COVID19 patients, demonstrating acute peripheral activation (Fig 1B). We next analysed neutrophil degranulation and detected increased surface abundance of CD66b, a secondary granule marker (44), across all patient groups (Fig 1C). We also detected a trend towards increased abundance of CD63 (primary granule marker) positive neutrophils, in severe patients (Sup Fig 2E), but found no difference in CD11b expression (tertiary and secretory vesicle marker) (Sup Fig 2F). CD66b exocytosis and loss of CD62L expression did not differ with survival status of severe patients (Sup Fig 2G).

To further explore secondary granule release, we analysed expression of CD177, a protein that localises to both the plasma membrane and secondary granules and is upregulated upon degranulation (45). In the majority of healthy individuals, CD177 demonstrates bimodal expression, with approximately 50% of circulating neutrophils expressing this receptor (46) (Sup Fig 2H). We detected significant upregulation of CD177 expression in neutrophils from moderate and severe COVID-19 groups (Fig 1D), and a significant increase in overall percentages of CD177^+^ neutrophils (Sup Fig 2H). Furthermore, patients who later succumbed to disease displayed a trend towards greatest expression of CD177 (Sup Fig 2I). In summary, moderate and severe COVID-19 is associated with acute neutrophil activation and release of secondary granules.

### Severe COVID-19 patients maintain naïve CXCR2 expression levels

Trafficking of neutrophils to organs, including the lung, is orchestrated by chemokines such as CXCL8 (IL-8) and CXCL1 (47), which primarily signal via the CXCR2 chemokine receptor. This G protein-coupled receptor (GPCR) shapes the neutrophil chemotactic response by regulating the actin cytoskeleton (48). At high concentrations of chemokines, CXCR2 is internalised by endocytosis, which has been suggested as a form of terminal activation, after which cells are no longer responsive to cognate chemokines (49). Interestingly, CXCR2 expression was significantly reduced in moderately ill patients (Fig 1E). Severely ill patients, in contrast, maintained CXCR2 expression at levels similar to those found in healthy or convalescent individuals (Fig 1E), although there was no significant difference between survivors and non-survivors (Sup Fig 2J). We found a similar but non-significant trend towards maintenance of CXCR4 expression in severe patients (Sup Fig 2K). In summary, neutrophils in patients with moderate disease downregulate CXCR2 expression, potentially limiting their capacity to migrate to inflamed tissues. In contrast, neutrophils in patients with severe disease are characterised by a failure to downregulate CXCR2 expression, indicating maintained capacity for trafficking to organs such as the lungs.

### Neutrophils from patients with severe COVID-19 upregulate pathways for chemotaxis and protein synthesis

To further explore neutrophil activation and chemotactic capacity, we compared the proteomes of circulating neutrophils from severe COVID-19 patients (n=3; % CD10^−^ = 9.4 - 17.6%) and healthy controls (n=4). We detected a total of 3316 proteins (Fig 2A), including 466 proteins with significantly altered expression (*p*<0.05) (Fig 2B), of which 75 also passed a threshold of 2 fold difference. Principle component analysis (PCA) demonstrated heterogeneity amongst the severe patients, which was explained by patient outcome (Fig 2C); however due to the small sample size, subsequent analysis was performed on combined patient samples.

**Figure 2:**
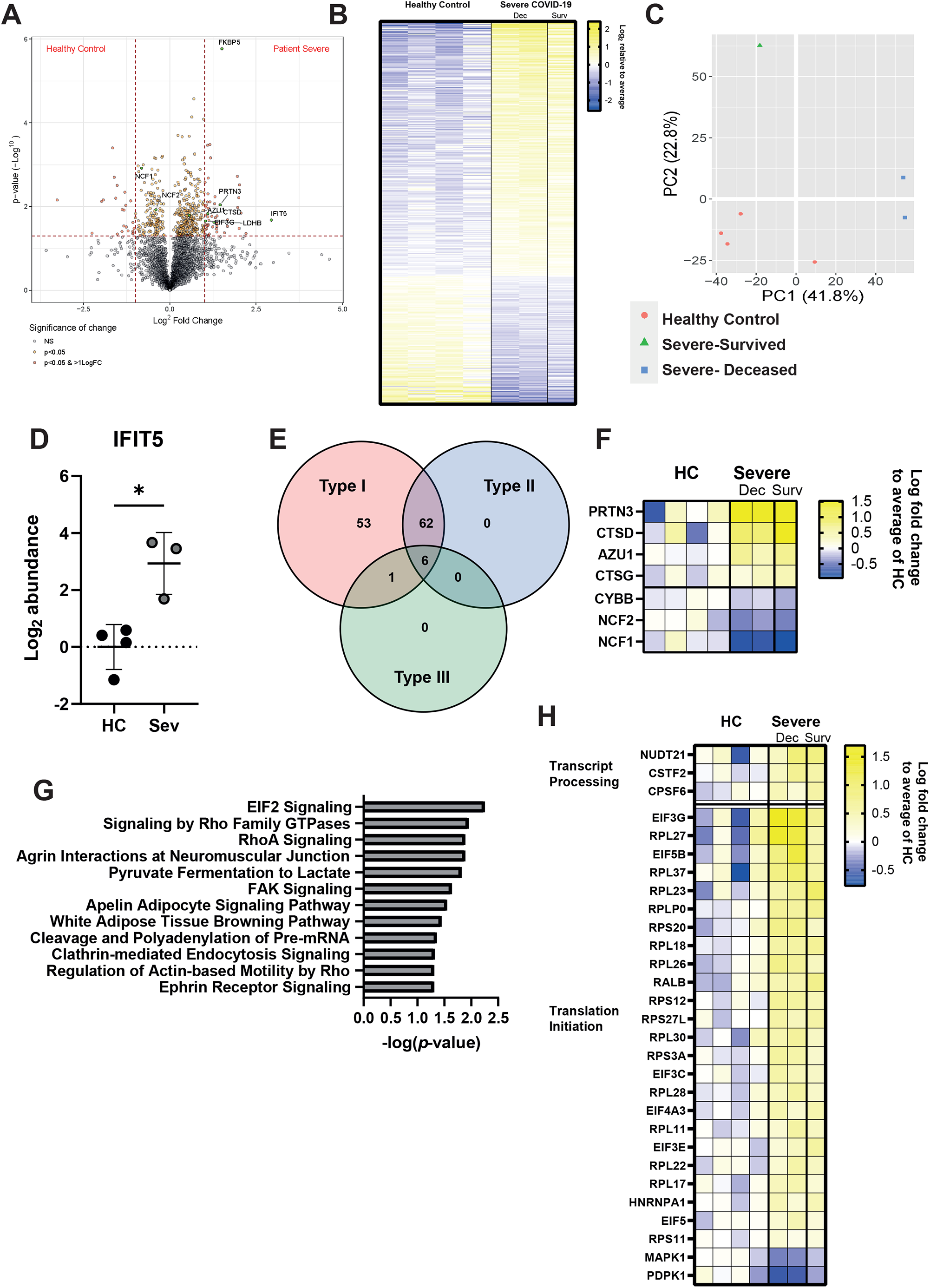
Severe COVID-19 induces numerous changes to the neutrophil proteome. A) Volcano plot comparing protein abundances between severe COVID-19 patients and healthy controls displayed by-log_10_ p-value and log_2_ fold change. B) Log_2_ abundance of proteins identified with significantly altered expression between severe COVID-19 and healthy controls relative to mean expression. C) PCA analysis of neutrophil proteomes, with distribution of healthy controls (red), severe COVID-19/survived (green) and severe/deceased (blue) indicated. D) Significantly altered proteins annotated by relation to interferon class signalling using the interferome database. E) Significantly altered proteins related to neutrophil function displayed as log_2_ fold change over healthy controls. F) IPA identified canonical pathways significantly enriched in severe COVID-19 patients neutrophils as ranked by -log_10_ *p*-value. G) Significantly altered proteins related to mRNA processing and EIF2 signalling as identified by IPA, displayed as log_2_ fold change over healthy controls. Statistical analysis for E was performed by Welch’s t test. Values represent mean ± standard deviation.

The most strongly induced significant protein was interferon-induced protein with tetratricopeptide repeats (IFIT) 5 (fold change= 7.65, p= 0.021) (Fig 2A and 2D), consistent with previous reports of a type I interferon signature in neutrophils from COVID-19 patients (50-53). Indeed, interrogation of significantly altered proteins using the Interferome database (54) confirmed active type I IFN signalling in severe COVID-19 patients, despite dexamethasone administration (Fig 2E).

We identified changes in numerous proteins related to neutrophil function. These included reductions in components of the NADPH oxidase complex (neutrophil cytosol factor [NCF 1 and 2] and cytochrome b 558 subunit beta [CYBB]) (Fig 2F), suggesting reduced ability to produce ROS and potentially contributing to the increased incidence of fungal and bacterial co-infections in steroid-treated COVID-19 patients (55). Conversely, proteins associated with neutrophil granules such as Azurocidin (AZU1) Cathepsin (CTS) D and G and proteinase (PRTN) 3 were elevated in patients, suggesting neutrophils may instead be attuned to hyperdegranulation (Fig 2F).

Ingenuity pathway analysis (IPA) identified pathways related to chemotaxis, such as ‘Signalling by Rho family GTPases’, ‘RhoA signalling’ and ‘Regulation of Actin-based Motility by Rho’ as significantly enriched in patient neutrophils (Fig 2G), which aligns with maintained expression of CXCR2 (Fig 1G). IPA also detected enrichment of RNA processing and translation pathways (‘Cleavage and polyadenylation of pre-mRNA’ and ‘eIF2 signalling’ (Fig 2G and 2H), suggesting elevated *de novo* protein synthesis.

In summary, proteomic changes in patient neutrophils highlight increased chemotaxis and protein translation, which is consistent with detection of activation by flow cytometry.

### CD10^−^ immature neutrophils are hyperactivated

We next used flow cytometry data to i) ask whether the highly enriched immature neutrophils differ phenotypically from their mature counterparts, and ii) identify subpopulations that are biomarkers of severe disease. We combined data from healthy controls, convalescent and COVID-19 patients and performed uniform manifold approximation and projection (UMAP) (Fig 3A), a form of dimensionality reduction that generates 2-dimensional representations of multiple surface markers and allows for interrogation of cellular heterogeneity (56). Distribution of healthy controls, convalescent and COVID-19 patients within the UMAP plot demonstrated that these three groups occupy relatively distinct spaces (Fig 3B). Interestingly, neutrophils from convalescent patients (median days since symptom onset =76.5±30.7) were characterised by an intermediate distribution between healthy controls and acute patients, suggesting long term perturbations to the myeloid compartment.

**Figure 3:**
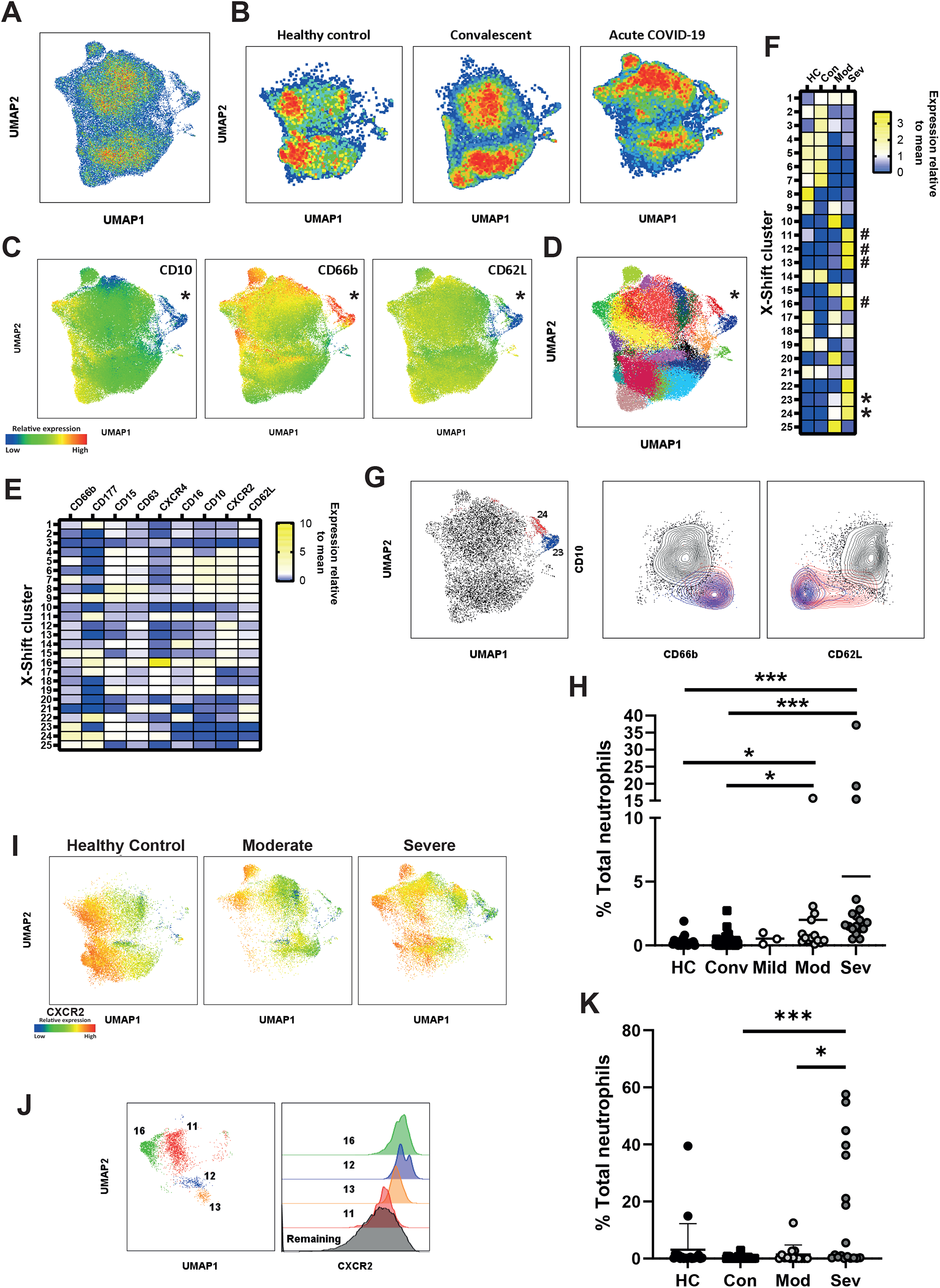
Hyperactive immature neutrophils are a feature of the immune response in severe COVID-19. A) UMAP clustering of circulating neutrophils combined from 90 individual donors. B) Pseudo-colour plots demonstrate distribution of healthy controls and convalescent donors and acute COVID-19 patients within UMAP plot. C) Expression of CD10, CD66b and CD62L overlayed on UMAP plot with population of interest marked with an asterisk. D) X-shift clusters overlayed onto UMAP plots. F) Relative distribution of X-Shift clusters between stratified groups. * hyperactive immature clusters; # CXCR2 high clusters. G) X-shift clusters 24 (red) and 23 (blue) displayed on UMAP plot (left), and density plots (right) comparing clusters 24 and 23 to remaining clusters for CD10 against CD66b and CD62L. H) Distribution of clusters 23 and 24 neutrophils among stratified groups. I) UMAP plots stratified into healthy control and patient neutrophils with expression of CXCR2 overlayed. J) X-shift clusters 11, 12, 13 and 16 as displayed on UMAP plot (left), histogram demonstrating CXCR2 expression in clusters 11, 12, 13, 16 and neutrophils from remaining clusters (right). K) Distribution of neutrophils from X-shift clusters 11, 12, 13 and 16 among stratified groups. G and J were analysed by Kruskal-Wallis test with Dunn’s multiple comparisons displayed on graph. Values represent mean ± standard deviation.

**Figure 4:**
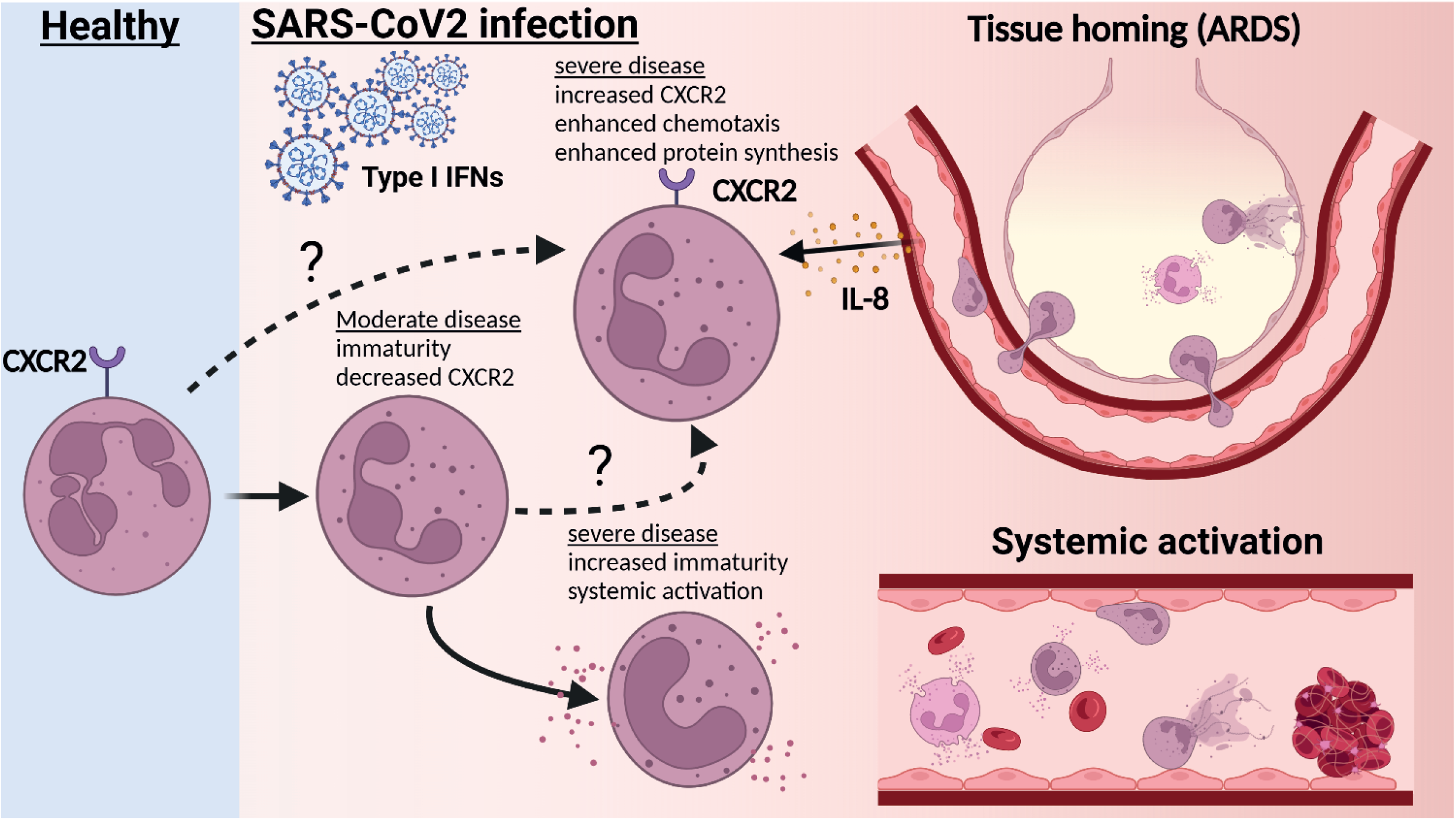
Severe patient neutrophils fail to engage in protective reprogramming and instead engage in systemic activation. Upon acute infection with SARS-CoV2, peripheral neutrophils become increasingly immature with reductions in markers associated with maturity, such as CD10. In moderate patients this is accompanied by reductions in CXCR2 expression. In severe patients, CXCR2 is maintained in a subset of neutrophils. This potentially aids recruitment to infected lung tissue and may contribute to lung injury. Additionally, CD10^−^ immature neutrophils in severe patients become activated in the periphery, with increased markers of degranulation and surface protease activity. These immature clusters lack CXCR2 expression, suggesting that instead of trafficking to inflamed organs, they may contribute to peripheral pathology, such as immunthrombosis.

To examine the functional state of immature neutrophils, we overlayed surface marker expression on UMAP plots, which demonstrated distinct, alternatively activated clusters (Fig 3C). CD10^−^ cells were identified as a discrete population (marked by asterisk), that was simultaneously CD66b^high^ and CD62L^low^ (Fig 3C). This cluster was also identified as CXCR2^low^, suggesting it does not account for maintenance of CXCR2 in severe patients (Sup Fig 3A). We conclude that peripheral CD10^−^ cells are hyperactive, engaging in increased secondary granule exocytosis and surface protease activation.

To validate our UMAP clustering, and to unbiasedly identify clusters associated with severe disease, we combined UMAP clustering with X-shift analysis (57), an algorithm which automatically identifies cell populations and selects the optimal number of clusters. Using this approach, 25 clusters were identified, which were then overlayed onto UMAP plots (Fig 3D and E). Distribution of X-Shift clusters between stratified groups supports our observation of distinct neutrophil phenotypes in different disease states (Fig 3F). This approach also confirmed our manual gating: ranking clusters by CD10 expression identified the CD10^−^ immature clusters (Fig 3C), as two clusters (clusters 23 and 24), separated by the bimodal expression of CD177 (Fig 3G and Sup Fig 3B). Examination of surface expression in these clusters identifies these as possessing the greatest CD66b expression and the lowest CD62L expression among all detected clusters (Fig 3E and G). These clusters were significantly enriched in severe and moderate patients, demonstrating they are a feature of worsening disease (Fig 3H). Taken together, our analysis identifies hyperactive CD10^−^ neutrophil subpopulations as potential biomarkers of severe COVID-19.

### CXCR2 expressing neutrophils are associated with severe COVID-19

UMAP analysis confirmed the maintenance of CXCR2 expressing neutrophils in severe disease (Fig 3I). Interestingly, X-shift analysis identified additional clusters specific for severe COVID-19 (clusters 11, 12, 13 and 16) (Fig 3F and Sup Fig 3C). These clusters all showed elevated expression of CXCR2 (Fig 3J) and were notably absent in moderately ill patients (Fig 3K, Sup Fig 3C). On the other hand, the clusters were significantly elevated in severe disease (Fig 3K) and may therefore represent useful prognostic biomarkers. Such enrichment of CXCR2^hi^ clusters suggest that neutrophils from severe patients may maintain responsiveness to chemokines such as IL-8.

### Partially activated immature neutrophils persist 3 months after infection

We next asked whether there are any unique clusters associated with convalescence, as suggested by UMAP analysis (Figure 3B). We identified a single subpopulation (cluster 3) which was enriched in convalescent patients (Fig 3F and Sup Fig 3D). Interestingly, cluster 3 also showed reduced CD10 expression (Sup Fig 3E), indicating that immature neutrophils persist up to 3-months post infection. This convalescent-enriched cluster was not associated with degranulation (CD66b low) but did demonstrate partial activation, as evident from CD62L shedding (Sup Fig 3E). When we stratified convalescent individuals into either outpatients or hospitalised patients, we found cluster 3 to be significantly enriched in individuals who had been admitted to hospital due to COVID-19 complications (Sup Fig 3F and 3G). This finding demonstrates that neutrophil dysregulation persists for prolonged periods after viral clearance.

## Discussion

Neutrophils were historically considered to be homogeneous cells, but recent studies have highlighted their phenotypic and functional plasticity (58). We hypothesized that divergent neutrophil developmental states may be important determinants of COVID-19, a disease that is characterised by a disordered myeloid response and a hyperinflammatory clinical phenotype (42, 59, 60).

Our study confirms multiple reports of expansion of neutrophil precursors in COVID-19 and their association with severe disease (32, 34, 61, 62). Using multiparameter cytometric analyses, we show that CD10^−^ neutrophils engage in hyper-degranulation of secondary granules, as well as proteolytic cleavage of the surface receptor CD62L (42). Consistent with our results, MMP8 and other secondary granule components are significantly elevated in plasma of ICU versus non-ICU COVID-19 patients (63). Furthermore, multiple studies have highlighted CD177, a protein associated with secondary granules, as predictive of poor clinical outcome (64-66). Secondary granules contain multiple proteins involved in cytoadhesion (67, 68), as well as metalloproteases with immunomodulatory and tissue degradation properties (69, 70), suggesting possible enhanced endothelial adhesion by degranulated CD10^low^ neutrophils. Furthermore, the contents of these granules also mediate interactions with platelets (71). Circulating CD10^−^ neutrophils may thus specifically be related to immunothrombus formation and coagulopathy (72).

Our findings are consistent with other reports of altered function of immature neutrophils. Circulating CD10^low^ neutrophils are thought to result from premature release from the bone marrow and have been detected in multiple diseases, including cancer (73) and bacterial sepsis (74). They are mobilised by the growth factor GCSF (75), which is elevated in COVID-19 patients (19). CD10^−^ neutrophils typically sediment with ‘low density granulocytes’ and PBMCs on Ficoll density gradients (38, 76). In vasculitis, CD10^−^ neutrophils have reduced ability to generate NETs but higher capacity to cause endothelial permeability, compared to CD10^+^ neutrophils (77). Similarly, in systemic lupus erythematosus, immature CD10^−^ neutrophils were found to have reduced capacity to phagocytose bacteria and produce NETs but had elevated propensity to degranulate (76). In lung cancer, CD10^low^ neutrophils correlate with advanced disease and also co-express PD-L1, raising the possibility that immature neutrophils may also have suppressive functions (78). These studies illustrate the need for deep phenotypic characterisation of CD10^−^ cells, with directed mechanistic studies, to fully understand how this developmental shift impacts disease progression.

In contrast to CD10 expression, which was reduced on all neutrophils from COVID-19 patients, we detected severity-specific differences in CXCR2 expression. Moderate disease was accompanied by reduced CXCR2 surface abundance, while expression was maintained at baseline levels in severe patients. CXCR2 is known to be dynamically regulated, both during maturation and inflammation. Typically, CXCR2 expression is gradually increased during maturation (79) but can also be rapidly downregulated by internalisation upon exposure to high concentrations of ligand (49). Since both moderate and severe COVID-19 associated neutrophils are immature, it is unlikely that a difference in maturation state between these groups explains the divergent expression of CXCR2. In mouse models of influenza, CXCR2 is required for homing of neutrophils to the lung (80), and treatment with a CXCR2 inhibitor reduced lung pathology (81). Downregulation of CXCR2 in moderate COVID-19 may therefore be a protective, adaptive response that fails to occur in severe disease. Maintenance of neutrophil CXCR2 expression in severe COVID-19 may promote lung trafficking and associated tissue damage and perfusion defects. The role of IL-8/CXCR2 signalling in disease progression is supported by whole blood RNA expression profiling (82) and an independent proteomic study (50), both of which report specific upregulation of the pathway in severe COVID-19. CXCR2 inhibitors may have value as therapeutic agents in severe COVID-19 (83).

Consistent with a role for CXCR2 signalling, our mass spectrometry results revealed upregulation of pathways implicated in cell motility and protein synthesis, in addition to a strong type I interferon response. Our proteomic analysis also identified signatures of protein synthesis in neutrophils from severe COVID-19 patients, namely EIF2 signalling. Others have identified upregulation of the related EIF4 pathway, similarly involved in protein translation, in severe COVID neutrophils (41). These findings suggest that neutrophil translational activity may be contributing to immunopathogenesis.

A limitation of our study was the sample size where, particularly for proteomics, analysis of data following stratification by patient outcome was unfortunately not possible or underpowered. This risks the reduced detection of significant alterations to protein content due to the low number of patient samples.

In summary, we show that severe COVID-19 is associated with hyperactive immature neutrophils, maintenance of CXCR2 expression and increased translational activity. Our study identifies neutrophil subpopulations as prognostic biomarkers of disease severity and supports a role for neutrophils in COVID-19 immunopathogenesis. CXCR2 downregulation on neutrophils may act to prevent progression to severe disease. Finally, as proposed by others, the IL-8 signalling pathway, and in particular CXCR2, may represent therapeutic targets for severe COVID-19 (84).

## Materials and methods

### Human subjects and samples

Written informed consent was obtained from all patient and healthy donors, or from patients’ family if patients were too unwell to consent. Samples were obtained under research ethics approval of the DISCOVER study (Diagnostic and Severity markers of COVID-19 to Enable Rapid triage, NHS REC 20/YH/0121) and Bristol BioBank (NHS REC 20/WA/0053). The convalescent patient cohort consisted of a mixture of recovered outpatient healthcare workers, recruited through Bristol BioBank, and DISCOVER patients, who were recalled for follow up 3 months after initial hospital admission. Donor demographics and clinical data are described in Table 1. Patients were stratified by disease severity as described in Arnold *et al*. (35). Venous blood was collected in EDTA tubes (BD Biosciences).

### Flow cytometry

**S**taining was performed on whole blood. 100μl of blood was washed in PBS and incubated with Zombie Aqua live/dead stain (biolegend, product number 423101) at room temperature for 10 minutes. Samples were incubated with FC-Block (Biolegend, Human TruStain FcX™, product number 422302) in staining buffer (PBS, 5mM EDTA, 0.5%BSA), followed by addition of primary antibodies in staining buffer and incubation on ice for 20 minutes. Cells were washed and incubated with fluorophore conjugated streptavidin on ice for 15 minutes. Erythrocytes were lysed with chilled ACK lysis buffer (Gibco, product number A10492-01) and cells were fixed in 4% PFA for 20 minutes. Cells were resuspended in staining buffer and analysed on a BD X20 Fortessa flow cytometer. Data were analysed using FlowJo (FlowJo, LLC). See Supplementary Table 1 for flow cytometry reagents.

### UMAP and flowsom analysis

Dimensionality reduction was performed using concatenated live neutrophil populations from all stratified groups using 1000 events per donor (see supplementary data for gating strategy). UMAP plugin was used on the flowjo software platform to generate dimensionality reduction plots using neutrophil surface markers CD66b, CD177, CD15, CD63, CXCR4, CD16, CD10 CXCR2 and CD62L and the following setting; Euclidean 15, nearest neighbours 15, minimum distance 0.5 and number of components 2. X-Shift clustering was generated using the flowjo platform and used the same markers as UMAP, with the following settings: number of nearest neighbours (k): 20, Distance Metric: Angular, sampling limit 94000. Flowsom was generated using the flowjo platform and used the same markers as UMAP and was set for 25 and 8 meta clusters.

### Proteomic analysis

Protein lysates were obtained from neutrophils isolated with Stem Cell whole blood neutrophil isolation kit (Stemcell) as per manufacturer’s instructions. 5×10^6^ cells were washed in PBS and resuspended in RIPA buffer supplemented with 2x Protease inhibitor cocktail (Calbiochem), 2mM PMSF (Sigma), 1x Halt Phosphatase (Thermo Scientific), 50mM TCEP (Sigma) and 10mM EDTA. To limit potential biohazard risk, samples were heat inactivated at 57°C for 30 min prior to storing at -20°C. Protein was quantified using Pierce BCA™ protein assay kit (Thermo scientific) and 100 μg of protein used for Tandem-Mass-Tag (TMT) mass spectrometry.

50µg of each sample was digested with trypsin (1.25µg; 37°C, overnight), labelled with Tandem Mass Tag (TMT) eleven plex reagents according to the manufacturer’s protocol (Thermo Fisher) and the labelled samples pooled. A 100µg aliquot of the pooled sample was evaporated to dryness, resuspended in 5% formic acid and then desalted using a SepPak cartridge according to the manufacturer’s instructions (Waters). Eluate from the SepPak cartridge was again evaporated to dryness and resuspended in buffer A (20 mM ammonium hydroxide, pH 10) prior to fractionation by high pH reversed-phase chromatography using an Ultimate 3000 liquid chromatography system (Thermo Scientific). In brief, the sample was loaded onto an XBridge BEH C18 Column (130Å, 3.5 µm, 2.1 mm X 150 mm, Waters) in buffer A and peptides eluted with an increasing gradient of buffer B (20 mM Ammonium Hydroxide in acetonitrile, pH 10) from 0-95% over 60 minutes. The resulting fractions (15 in total) were evaporated to dryness and resuspended in 1% formic acid prior to analysis by nano-LC MSMS using an Orbitrap Fusion Tribrid mass spectrometer (Thermo Scientific).

High pH RP fractions were further fractionated using an Ultimate 3000 nano-LC system in line with an Orbitrap Fusion Tribrid mass spectrometer. In brief, peptides in 1% (vol/vol) formic acid were injected onto an Acclaim PepMap C18 nano-trap column (Thermo Scientific). After washing with 0.5% (vol/vol) acetonitrile 0.1% (vol/vol) formic acid peptides were resolved on a 250 mm × 75 μm Acclaim PepMap C18 reverse phase analytical column (Thermo Scientific) over a 150 min organic gradient, using 7 gradient segments (1-6% solvent B over 1min., 6-15% B over 58min., 15-32%B over 58min., 32-40%B over 5min., 40-90%B over 1min., held at 90%B for 6min and then reduced to 1%B over 1min.) with a flow rate of 300 nl min^−1^. Solvent A was 0.1% formic acid and Solvent B was aqueous 80% acetonitrile in 0.1% formic acid. Peptides were ionized by nano-electrospray ionization at 2.0kV using a stainless-steel emitter with an internal diameter of 30 μm (Thermo Scientific) and a capillary temperature of 275°C. All spectra were acquired using an Orbitrap Fusion Tribrid mass spectrometer controlled by Xcalibur 2.1 software (Thermo Scientific) and operated in data-dependent acquisition mode using an SPS-MS3 workflow. FTMS1 spectra were collected at a resolution of 120 000, with an automatic gain control (AGC) target of 200 000 and a max injection time of 50ms. Precursors were filtered with an intensity threshold of 5000, according to charge state (to include charge states 2-7) and with monoisotopic peak determination set to peptide. Previously interrogated precursors were excluded using a dynamic window (60s +/-10ppm). The MS2 precursors were isolated with a quadrupole isolation window of 1.2m/z. ITMS2 spectra were collected with an AGC target of 10 000, max injection time of 70ms and CID collision energy of 35%.

For FTMS3 analysis, the Orbitrap was operated at 50 000 resolution with an AGC target of 50 000 and a max injection time of 105ms. Precursors were fragmented by high energy collision dissociation (HCD) at a normalised collision energy of 60% to ensure maximal TMT reporter ion yield. Synchronous Precursor Selection (SPS) was enabled to include up to 10 MS2 fragment ions in the FTMS3 scan.

The raw data files were processed and quantified using Proteome Discoverer software v2.1 (Thermo Scientific) and searched against the UniProt Human database (downloaded January 2021) using the SEQUEST HT algorithm. Peptide precursor mass tolerance was set at 10ppm, and MS/MS tolerance was set at 0.6Da. Search criteria included oxidation of methionine (+15.995Da), acetylation of the protein N-terminus (+42.011Da) and Methionine loss plus acetylation of the protein N-terminus (−89.03Da) as variable modifications and carbamidomethylation of cysteine (+57.021Da) and the addition of the TMT mass tag (+229.163Da) to peptide N-termini and lysine as fixed modifications. Searches were performed with full tryptic digestion and a maximum of 2 missed cleavages were allowed. The reverse database search option was enabled and all data was filtered to satisfy false discovery rate (FDR) of 5%.

### Bioinformatics analysis of Proteomics

Following analysis in Proteome Discoverer 2.1, the proteomics data were processed and further analysed in the R statistical computing environment. The protein groups were reassessed by an in-house script which selects a master protein firstly by ID and quantitation metrics, then by the annotation quality of uniprot accessions. Data were log2 transformed, and statistical significance calculated using Welch’s t test. IPA analysis was performed with a filter of P<.05 to identify biological trends in the proteins which are statistically significant between clinical conditions. PCAs were calculated using the PCA function in the FactoMineR package, and plotted using either ggplot (2D), or Plotly (3D).

### Statistical analysis

Statistical analysis was performed using Graphpad Prism 8. Where appropriate, MFI data were log transformed prior to analysis by one way ANOVA or student t test * *p*<0.05, ** *p*<0.01, *** *p*<0.001

## Data Availability

All data produced in the present study are available upon reasonable request to the authors

## Author contributions

**CMR** conceived the study, performed experimental procedures, analysed data and wrote the manuscript; **PL** analysed data; **FPG, WG, DC, AD** performed experiments and analysed data; **FH, DA, AG, CH, LO, RB, AF** and **LW** conceived the clinical aspects of the study and organised sample collection; **LR** helped conceive the study; **BA** conceived and supervised the study and wrote the manuscript. All authors read, provided input and approved the final manuscript.

## Acknowledgments

We thank Andrew Herman and Helen Rice for assistance with flow cytometry and Kate Heesom for proteomics. We also thank Marianna Santopaolo, Michaela Gregorova and Lea Knezevic for help with sample processing, as well as Jill King, Jane Metz, Charlie Plumptre, Begonia Morales-Aza, Lucy Collingwood and Jenny Oliver for their work in assisting with participant recruitment and consent as part of Bristol Biobank.

## Figure legends

**Supplemental Figure 1.**
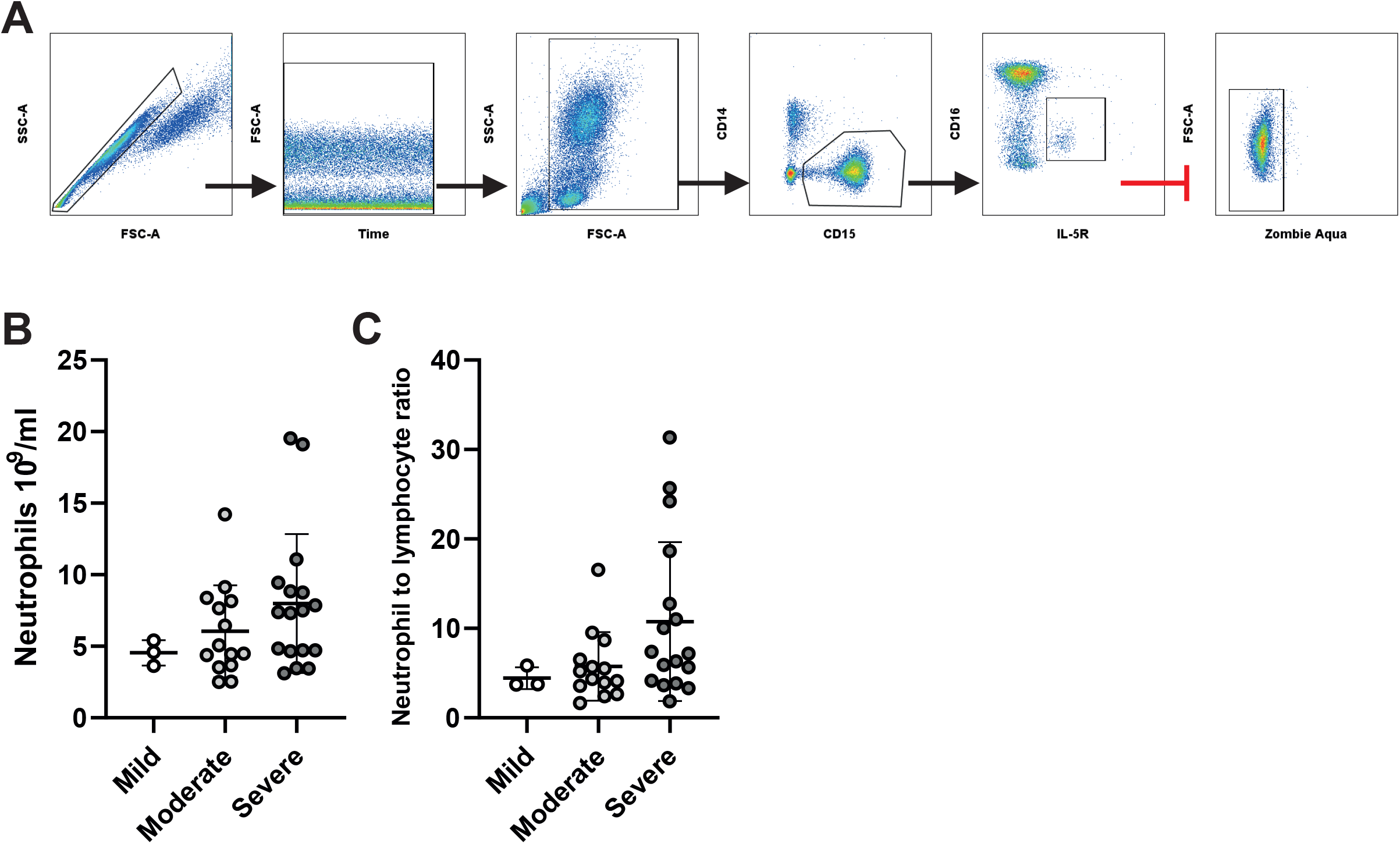
a) Gating strategy for peripheral blood neutrophil analysis. Single cells were discerned using forward to side scatter ratio, time gates were used to eliminate any inconsistencies in acquisition, debris elimination gates removed low forward and side scatter events, CD15^+^/CD14^-^ granulocytes gated and eosinophils were eliminated by expression of IL-5R (CD125). Finally dead neutrophils were removed by gating on zombie aqua negative cells. b) Peripheral blood neutrophil count between stratified patient groups. c) Neutrophil to lymphocyte ratio between stratified patient groups. Values represent mean ± standard deviation.

**Supplemental Figure 2.**
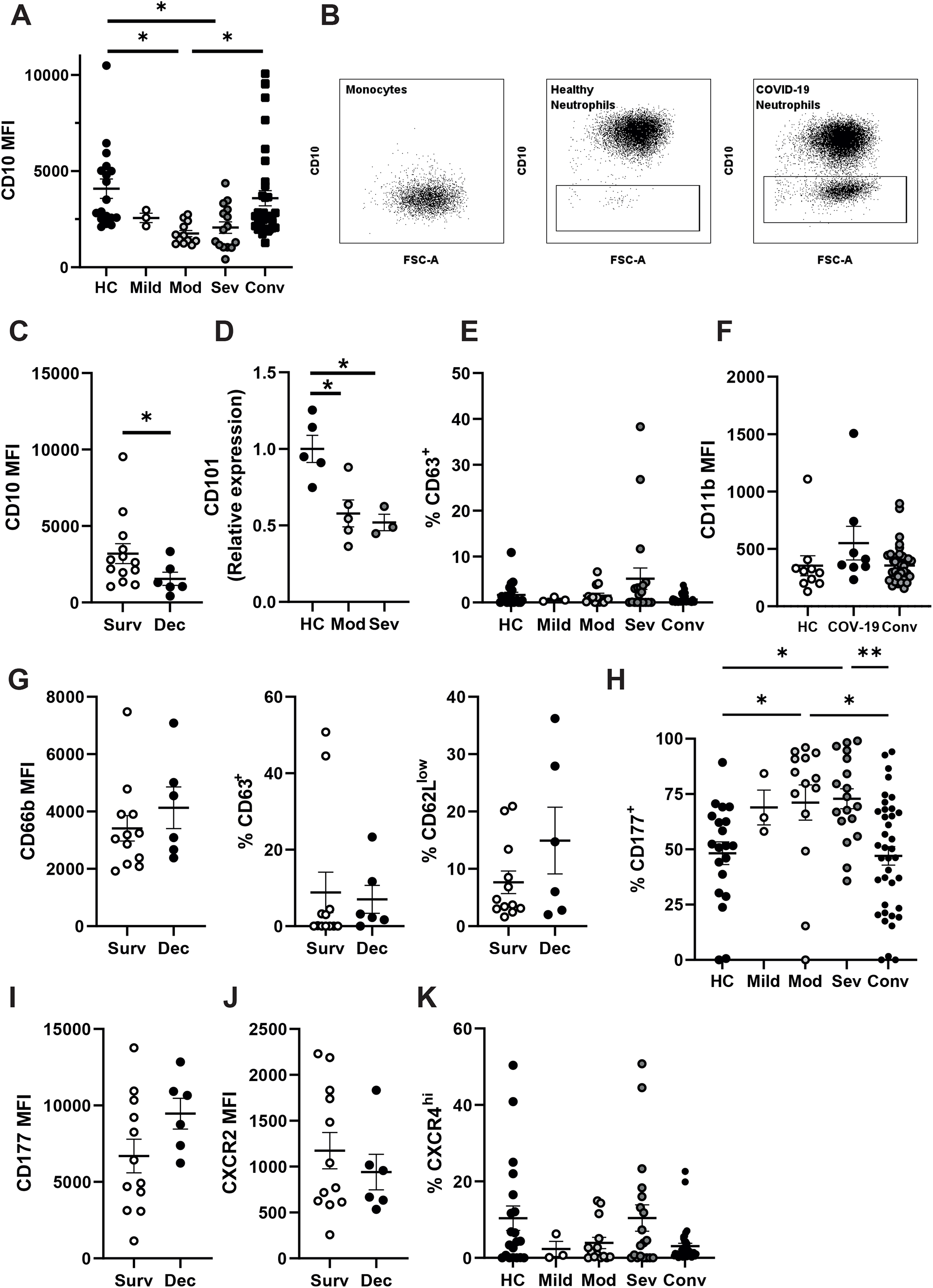
A) Neutrophil CD10 median fluorescent intensity from stratified groups. B) Representative gating for CD10^−^ neutrophils and comparison to CD10-monocytes. C) Neutrophil CD10 median fluorescent intensity from severe patients stratified by survival. D) Neutrophil CD101 median fluorescent intensity from healthy control, moderate and severe COVID-19 patients. D) Percentage of neutrophils expressing cell surface CD63 from stratified groups. E) CD11b median fluorescent intensity from Healthy control, acute COVID-19 patients and convalescent individuals. G) Neutrophil CD66b median fluorescent intensity, percentage of CD63^+^ and CD62L^low^ neutrophils from severe patients stratified by survival. H) percentage of CD177^+^ neutrophils from stratified groups. I) CD177 median fluorescent intensity of CD177^+^ neutrophil from severe patients stratified by survival. J) Neutrophil CXCR2 median fluorescent intensity from severe patients stratified by survival. K) Percentage of CXCR4^+^ neutrophils from stratified groups. Statistical analysis was performed by; A, data were log transformed and One-way ANOVAs with Tukey’s multiple comparisons displayed on the graph, C) data were log transformation and assessed by unpaired *t*-test, D) One-way ANOVAs with Tukey’s multiple comparisons displayed on the graph, H) was performed by Kruskal-Wallis test with Dunn’s multiple comparisons displayed on graph. Values represent mean ± standard deviation.

**Supplemental Figure 3.**
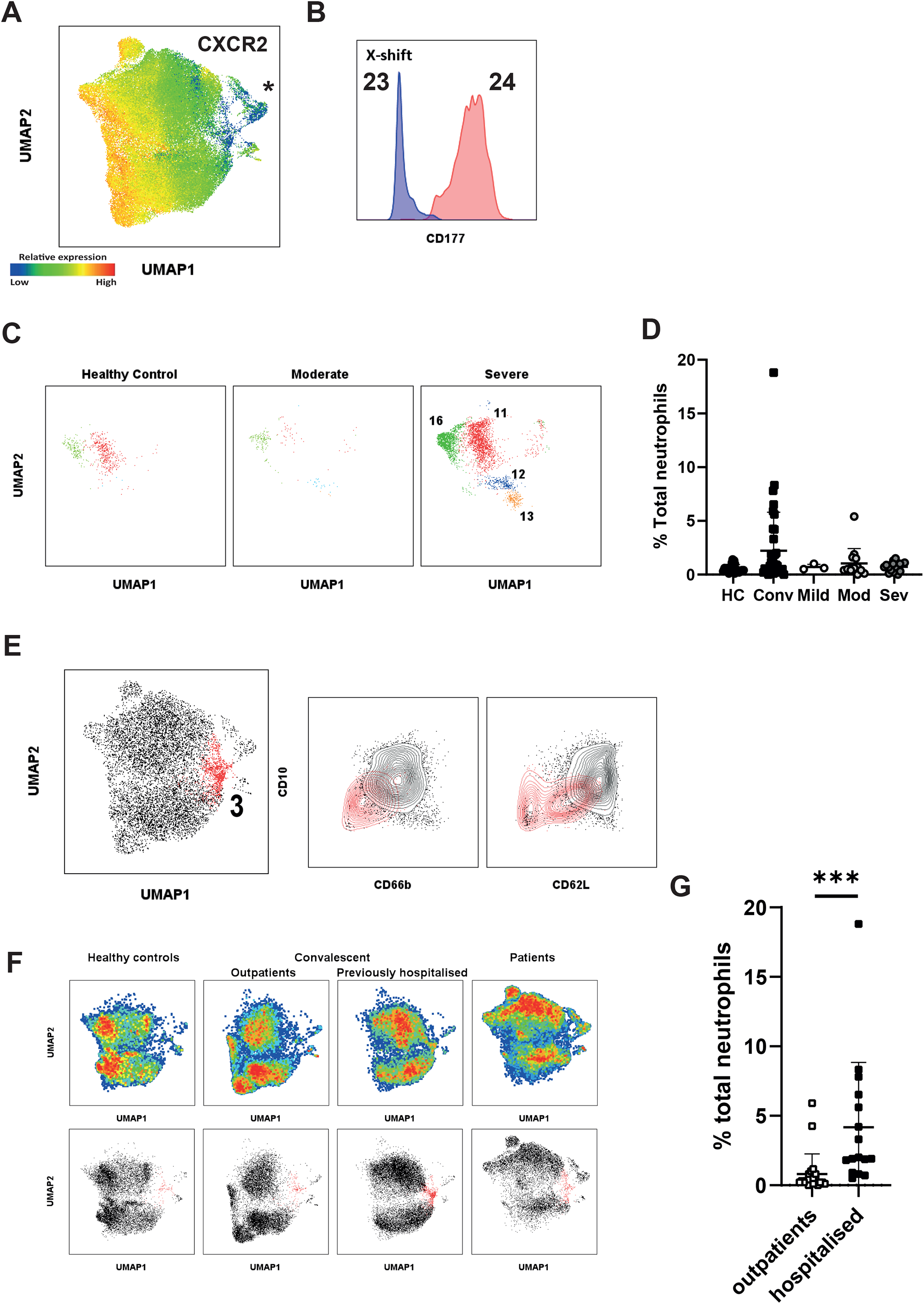
A) Relative expression of CXCR2 overlayed on UMAP plots of combined neutrophils, asterisk marks population of interest. B) Median fluorescent intensity of surface markers from 25 automatically generated X-shift clusters. C) CD177 expression between X-Shift clusters 23 and 24. D) X-Shift clusters 11, 12, 13 and 16 displayed on UMAP plots stratified between healthy control and moderate and severe COVID-19 patients. E) Distribution of neutrophils from X-shift clusters 3 among stratified groups. F) X-shift clusters 3 (red) displayed on U-MAP plot (left), and density plots (right) comparing cluster 3 to remaining clusters for CD10 against CD66b and CD62L. G) Pseudo-colour plots demonstrate distribution of healthy controls and convalescent outpatients, previously hospitalised convalescent and acute COVID-19 patients within U-MAP plot (top). Stratified UMAP plots with cluster 3 highlighted in red (bottom). H) Distribution of cluster 3 in convalescent individuals, stratified by hospitalisation status. Statistical analysis for; E) was performed by Kruskal-Wallis test with Dunn’s multiple comparisons displayed on graph, H) was performed by Mann Whitney test. Values represent mean ± standard deviation.

**Supplemental table 1.** Flow cytometry reagents used for study.

| Target-fluorophore | clone | company | Product code |
| --- | --- | --- | --- |
| CD63 Brilliant violet 421 | H5C6 | Biolegend | 353030 |
| CXCR4 Brilliant violet 605 | 12G5 |  | 306522 |
| CD14 Brilliant violet 785 | M5E2 |  | 301840 |
| CD16 FITC | 3G8 |  | 302006 |
| CD10 PE | HI10a |  | 312204 |
| CXCR2 PE | 5E8 |  | 320722 |
| CD62L PE Cy7 | DREG-56 |  | 304822 |
| CD15 Alexafluor 700 | HI98 |  | 301920 |
| CD66b APC | G10F5 |  | 305118 |
| CD177 APC-Cy7 | MEM-166 |  | 315810 |
| CD101 PER-CP Cy5.5 | BB27 |  | 331015 |
| Streptavidin PER-CP Cy5.5 | N/A |  | 405214 |
| Streptavidin 711 |  |  | 405241 |
| Zombie Aqua |  |  | 423101 |

